# Incidence of antidepressant withdrawal reactions: A prospective longitudinal cohort study in primary care patients

**DOI:** 10.64898/2026.05.12.26352975

**Authors:** Andri Rennwald, Mark A. Horowitz, Oliver Senn, Stefan Neuner-Jehle, Michael P. Hengartner

## Abstract

**Background:** The incidence of antidepressant withdrawal reactions in longer-term users and the influence of dosage is insufficiently understood.

**Objectives:** Informed by neuropharmacological models and user surveys, this study examined symptom change during tapering and if increases were specifically associated with reductions below 75% of the minimum effective dose.

**Design:** This was a prospective longitudinal cohort study with seven assessments over six months.

**Methods:** Altogether 32 Swiss adult primary care patients who were on antidepressants for at least six months and in stable remission were assessed at baseline (week 0) before they started tapering and after 2, 4, 6, 8, 16, and 26 weeks. Withdrawal symptoms were measured repeatedly using an adapted version of the Discontinuation-Emergent Signs and Symptoms Scale (DESS) and the main outcome was intra-individual symptom change during intervals. Antidepressant dose was standardized relative to the minimum effective dose in the treatment of depressive and anxiety disorders.

**Results:** Across intervals, reductions below 75% of the minimum effective dose were associated with symptom increases, while reductions above that threshold or no reductions were associated with symptom decreases. After adjusting for potential confounders, the rate of clinically relevant symptom increases contingent on dose reductions below 75% of the minimum effective dose was 33%, as compared to 13% during intervals with no dose reductions (OR=3.2, 1.4 to 7.4). We thus estimated that 60% of the risk of clinically relevant symptom increases was attributable to pharmacological withdrawal effects. The adjusted incidence rates for clinically relevant and severe withdrawal reactions were 32% and 11%, respectively.

**Conclusions:** Consistent with neuropharmacological research findings, we found that antidepressant withdrawal symptoms emerge mostly following reductions below 75% of the minimum effective dose, affecting about one-third of patients. Even small reductions may trigger clinically relevant withdrawal reactions in this lowest dose-range, stressing the need for personalized tapering plans.

## Introduction

Antidepressant prescribing has increased substantially in recent decades [1], largely due to growing long-term use [2]. Most people now stay on antidepressants for years, sometimes decades, often significantly longer than guidelines recommend [3,4]. While antidepressants are widely prescribed long-term, treatment discontinuation is often difficult, because patients may develop withdrawal symptoms that are distressing and impairing [5,6]. After decades of denial and minimisation [7,8], these problems have gained increased public and clinical attention in recent years, with media reports, patient advocacy and academic initiatives highlighting the issue and calling for more research [9–11].

Antidepressant withdrawal can affect different bodily and mental functions. Common physical symptoms include dizziness, imbalance and fatigue; salient emotional symptoms include irritability, mood instability and anxiety; and frequent cognitive symptoms comprise poor concentration and confusion [12,13]. Some patients experience mild and short-lived reactions, while others develop severe and more persistent withdrawal syndromes, but due to differences between drugs, unrepresentative samples and a lack of systematic longitudinal assessments, estimates are imprecise and uncertain [12,14]. While some research suggests that withdrawal reactions may affect more than half of patients, especially after long-term use [15], studies largely based on short-term treatment trials suggest incidence rates may be much smaller [16]. As a result, there is an ongoing controversy about the incidence and clinical relevance of antidepressant withdrawal reactions [17,18].

Although the Discontinuation-Emergent Signs and Symptoms Scale (DESS) has become the preferred research instrument to assess withdrawal reactions [17], the lack of specificity of most DESS items make it difficult to analyse and interpret its scores. Many symptoms included in the DESS not only overlap significantly with antidepressant side effects (e.g., sweating, trembling), but also with psychological stress (e.g., nervousness, trouble sleeping), frequent background ailments such as headaches or premenstrual symptoms (e.g., stomach cramps, irritability) and symptoms of mood and anxiety disorders (e.g., anxiety, mood swings). The comparably high mean DESS scores before start of tapering as evidenced in the REDUCE trial clearly demonstrate that the DESS captures more factors than just withdrawal effects [19]. This may result in an overestimation of incidence rates but also in under-detection of true-positive cases. For example, upon dose reduction, side effects may abate, while withdrawal symptoms emerge, thus offsetting each other and producing no apparent change in DESS score despite an incident withdrawal reaction. The sudden appearance or disappearance of external stressors and random symptom fluctuations due to general health problems may further complicate the detection of withdrawal reactions.

To delineate symptoms due to pharmacological withdrawal effects from symptoms attributable to other causes we thus chose to assess differences in intra-individual change in DESS scores contingent on current antidepressant dose change. Based on neuropharmacological research into serotonin transporter receptor occupancy in association with antidepressant dose, a pharmacological withdrawal effect is less likely to occur following dose reductions above the minimum effective dose but starts to manifest slowly below that threshold and will be most pronounced following dose reductions below three-fourths (75%) of the minimum effective dose [20,21]. By contrast, the occurrence of side effects, therapeutic effects and other factors, is not supposed to be restricted to a specific range below the minimum effective dose [22,23].

The aim of the present study is to advance our understanding of the risk of antidepressant withdrawal reactions by testing an empirically informed approach for assessing antidepressant withdrawal symptoms and to delineate pharmacological effects from other factors in a prospective longitudinal cohort study of primary care patients tapering their antidepressants.

## Methods

### Study design

This is a prospective observational cohort study over six months conducted as part of the AWARE project (Antidepressant Withdrawal in primary cARE patients). The study protocol was preregistered on the Open Science Framework (OSF) in November 2022; https://osf.io/aqmyt/overview. Adult primary care patients in stable remission planning to discontinue their antidepressant medication were referred to the research team by GPs and outpatient psychiatrists. Participants were interviewed immediately before starting the discontinuation process (baseline) and were re-assessed after 2, 4, 6, 8, 16, and 26 weeks. Comprehensive data on the tapering process, service use, withdrawal symptoms, mental health conditions and quality of life were collected throughout the six-month observation period. The study was conducted according to Strengthening the Reporting of Observational Studies in Epidemiology (STROBE) guidelines [24].

### Setting

Participants were recruited in the German-speaking part of Switzerland through general practitioners (GPs) and outpatient psychiatrists between November 2022 and October 2025. As specified in the study protocol, in November 2022 we first contacted 200 GPs who were randomly selected from the Swiss Medical Association (FMH) database of GPs in primary care. Since only four agreed to participate, another 200 general practitioners randomly selected from the FMH database were contacted by email in December 2022, yielding one additional response. Because five contributing GPs were deemed insufficient, further strategies were introduced in early 2023, including contacting regional healthcare services and outpatient psychiatrists (for a detailed description, see the supplement). Eventually, 10 GPs and four outpatient psychiatrists actively recruited study participants. To mitigate potential sources of bias, the term withdrawal symptoms was deliberately avoided in materials directed towards both the recruiting physicians and the study participants. Instead, the study was presented as a research project into the longer-term mental wellbeing and psychosocial functioning during and after discontinuing antidepressant medication.

### Participants

Eligible participants were adult primary care patients aged 18 to 75 years from the German-speaking part of Switzerland in stable remission who had taken antidepressants for at least six months and who were planning to discontinue their medication. They had to provide informed consent and be able to complete questionnaires in German. Exclusion criteria were insufficient knowledge of the German language, guideline-inconsistent short-term antidepressant use, persistent unremitted symptoms requiring ongoing treatment, and age above 75 years due to polypharmacy and multimorbidity complicating assessment of antidepressant withdrawal. Eligible participants were referred to the research team by their treating physicians and were enrolled into the study after confirmation of eligibility criteria and provision of informed consent. Once enrolled, participants remained in the study for 26 weeks. Assessments began before starting discontinuation of antidepressant medication (baseline, week 0) and continued prospectively through week 26. By the end of the recruitment period in October 2025, 33 patients had been referred by GPs and five by outpatient psychiatrists. Of these, six participants later withdrew from the study because they did not want to taper their antidepressant during the study period, leading to a final sample size of n=32.

### Variables and measurement

The primary outcome was the occurrence and severity of withdrawal symptoms, measured with an adapted version of the Discontinuation-Emergent Signs and Symptoms Scale (DESS) [25]. The DESS includes 43 withdrawal symptoms, which are assessed for worsening or new onset over the last two weeks. For the present study we added electroshock-like perceptions in the head (“brain zaps”) and removed the item “nose running” due to its high point prevalence in the Swiss population during the cold winter season. Following Michelson et al. [26] we rated symptoms as “absent”, “mild”, “moderate”, or “severe” instead of assessing worsening or new onset of symptoms as a dichotomous outcome (absent vs. present). The DESS was administered repeatedly at every measurement occasion to track changes in symptom presentation over time. Current antidepressant dose was reported by the participants at each measurement occasion in milligrams. To assess physical health and mental wellbeing at baseline we used the respective subscales from the short-form of the WHO Quality of Life questionnaire (WHOQOL-Bref; [27]). Demographic and clinical variables such as age, sex, treatment indication, and concomitant medication were also assessed at baseline. All data were collected through self-report using standardized online questionnaires in REDCap, a secure web-based platform. Baseline data were collected immediately before starting antidepressant discontinuation (week 0), partly by telephone (demographic and clinical variables) and partly by online questionnaire (outcome measures). All follow-up assessments were conducted online at weeks 2, 4, 6, 8, 16, and 26 after baseline.

### Study size

Due to unanticipated difficulties in the recruitment of study participants leading to significant delays in the data collection, we had to terminate the study prematurely before achieving our predefined sample size. Post-hoc power analysis focusing on repeated intra-individual changes up to week 26 (6 measurements per person) showed that with 32 participants we had in total 192 measurements, thus achieving a power of 0.83 to find a medium effect size of f=0.25 (for further explanations, see the supplement).

### Quantitative variables

In addition to the DESS total score we also calculated two DESS subscales informed by previous research [12,13], that is, neurosensory symptoms (unsteady gate/incoordination; dizziness; blurred vision; unusual visual sensation; burning/tingling sensations; unusual sensitivity to sound; ringing/noises in the ears; unusual tastes/smells, brain zaps) and affective symptoms (nervousness/anxiety; irritability; sudden mood worsening; sudden anger outbursts; sudden panic/anxiety attacks; bouts of crying/tearfulness; agitation; mood swings). DESS scores (0–3 for absent to severe per symptom) were calculated as sum scores at each time point and as intra-individual change scores from one time point to the next. Positive change scores thus indicate that symptoms have increased and negative change scores that they have decreased between subsequent time points. To assess the occurrence of a withdrawal syndrome we required the following three criteria: 1) the antidepressant dose had to be reduced from the previous dose, 2) the current dose had to be below three-fourth (75%) of the minimum effective dose, and 3) the DESS change score had to be positive. These criteria were chosen because an increase in DESS score is most likely caused by a pharmacological withdrawal effect when the dosage has been reduced in the subtherapeutic dose range [20,21]. An increase in DESS score was coded as subclinical or mild if the change score was positive but less than 4, as moderate if it was at least 4 points but less than 8, and as severe if it was 8 points or higher. One additional DESS change point indicates that one mild symptom has newly emerged or that one prevalent symptom has worsened from either mild to moderate or from moderate to severe. This categorization was chosen because it has been suggested that a clinically relevant withdrawal syndrome has to comprise at least 4 symptoms of any severity [28]. This definition also precludes overdetection and false-positive cases due to random symptom fluctuations and measurement error.

Current antidepressant dosage was standardized relative to each drug’s minimum effective dose in the treatment of depressive and anxiety disorders according to conventional prescribing guidelines in psychiatry [29]. For example, if a participant took 10mg of escitalopram, it was recorded as taking the minimum effective dose, if the dose was reduced to 7.5mg, it was recorded as 75% of the minimum effective dose, and so on. The antidepressants used were classified as SSRI and SNRI versus other drugs. Antidepressant treatment duration was extremely right skewed and was thus log-transformed. Finally, the antidepressant treatment indication was classified as an emotional disorder if participants reported being treated for a depressive or anxiety disorder.

### Statistical methods

All analyses were performed in SPSS version 31. We applied Generalized Estimating Equations (GEE), an extension of generalized linear models for the analysis of longitudinal data [30]. These models provide a parsimonious and flexible alternative to both random coefficient analysis and repeated-measures MANCOVA in longitudinal epidemiological research [31]. Following Twisk [32], we used a model-based estimator due to small sample size and the independent working correlation structure for the analysis of intra-individual change scores. In a preliminary analysis shown in the supplement, we tested whether we would find the assumed interaction effect between dose reduction relative to the minimum effective dose and intra-individual increases in DESS total scores. These models confirmed that dosage relative to 75% of the minimum effective dose (above vs. below) and dose change (reduction vs. no reduction) were statistically significant time-variant predictors both as main effects and in interaction. Further testing of different cut-offs confirmed that 75% of the minimum effective dose provided a better fit to the data than other thresholds (compared to 100% of the minimum effective dose and 125% of the minimum effective dose). As main predictor of repeated intra-individual changes in DESS score we thus used a three-factorial categorical variable comprising no dose reduction, reduction above 75% of minimum effective dose, and reduction below 75% of minimum effective dose (model 1). To adjust for major confounders at baseline, we conducted a separate model controlling for drug class, standardized dosage at baseline, treatment duration, and DESS baseline score (model 2). Finally, we also conducted a fully adjusted model which additionally controlled for age, gender, treatment indication, physical health at baseline, and mental wellbeing at baseline (model 3).

## Results

The sample is described in Table 1. At baseline the participants (66% women, mean age: 45.6 years) had been on their antidepressant on average for 42 months (SD=47.1), with a median of 29 months. The shortest duration of use was 6 months and the longest 168 months (14 years). Eight participants had attempted discontinuing their antidepressant before and failed. A specific tapering plan was provided to 12 participants (37.5%). Over half of the sample (56.3%) used escitalopram and standardized doses ranged from 50% to 250% of the minimum effective dose, with a mean of 140% (SD=58%). Four participants (12.5%) had a dose below the minimum effective dose before they started tapering.

**Table 1:** Description of the sample (n=32) at baseline (week 0)

| Continuous | Mean (SD) | Range |
| --- | --- | --- |
| Age (years) | 45.63 (17.19) | 18 to 75 |
| Duration of use (months) | 42.09 (47.07) | 6 to 168 |
| Standardized dose (%) | 140 (58) | 50 to 250 |
| Categorical | Option | N (%) |
| Gender | Man | 11 (34.4) |
|  | Woman | 21 (65.6) |
| Treatment Provider | GP | 27 (84.4) |
|  | Psychiatrist | 5 (15.6) |
| Previous discontinuation attempt failed | Yes | 8 (25.0) |
|  | No | 24 (75.0) |
| Antidepressant | Citalopram | 2 (6.3) |
|  | Escitalopram | 18 (56.3) |
|  | Sertraline | 3 (9.4) |
|  | Duloxetine | 1 (3.1) |
|  | Venlafaxine | 2 (6.3) |
|  | Agomelatine | 1 (3.1) |
|  | Bupropion | 1 (3.1) |
|  | Mirtazapine | 1 (3.1) |
|  | Trazodone | 1 (3.1) |
|  | Vortioxetine | 2 (6.3) |
| Treatment indication<br>(several responses possible) | Anxiety | 6 (18.8) |
|  | Depression | 17 (53.1) |
|  | Burnout | 6 (18.8) |
|  | Compulsions | 2 (6.3) |
|  | Trauma | 1 (3.1) |
|  | Stress | 2 (6.3) |
|  | Insomnia | 4 (12.5) |
|  | Pain | 1 (3.1) |
| Tapering plan recommended by<br>the treating physician | Abrupt (0 weeks) | 1 (3.1) |
|  | Fast (1-4 weeks) | 2 (6.3) |
|  | Standard (5-12 weeks) | 7 (21.9) |
|  | Slow (>12 weeks) | 2 (6.3) |
|  | No plan provided | 20 (62.5) |

Three participants (9.4%) stopped their drug abruptly and thus had their antidepressant discontinued by the second week. By week 8 (mean standardized dose: 48%, SD=48%), 15 participants (46.9%) had reduced their dose by up to 50% and 17 participants (53.1%) by more than 50%, of which 10 participants (31.3%) reduced their dose to zero. By week 16 (mean standardized dose: 32%, SD=44%), altogether 10 participants (31.3%) had no more than halved their initial dose, the others (68.7%) had reduced by more than 50%. By week 26 (mean standardized dose: 37%, SD=57%), altogether 15 participants (46.9%) had successfully discontinued their antidepressant, of which 12 had already discontinued their drug by week 16. As shown in Figure 1, the average dose reductions were larger at higher doses and became increasingly smaller when the dose went down. Above 75% of the minimum effective dose, the average dose reduction was −67% of the previous dose, while at doses below 75%, 50% and 25% of the minimum effective dose, the average dose reductions were −32%, −20%, and −12%, respectively of the previous dose.

**Figure 1:**
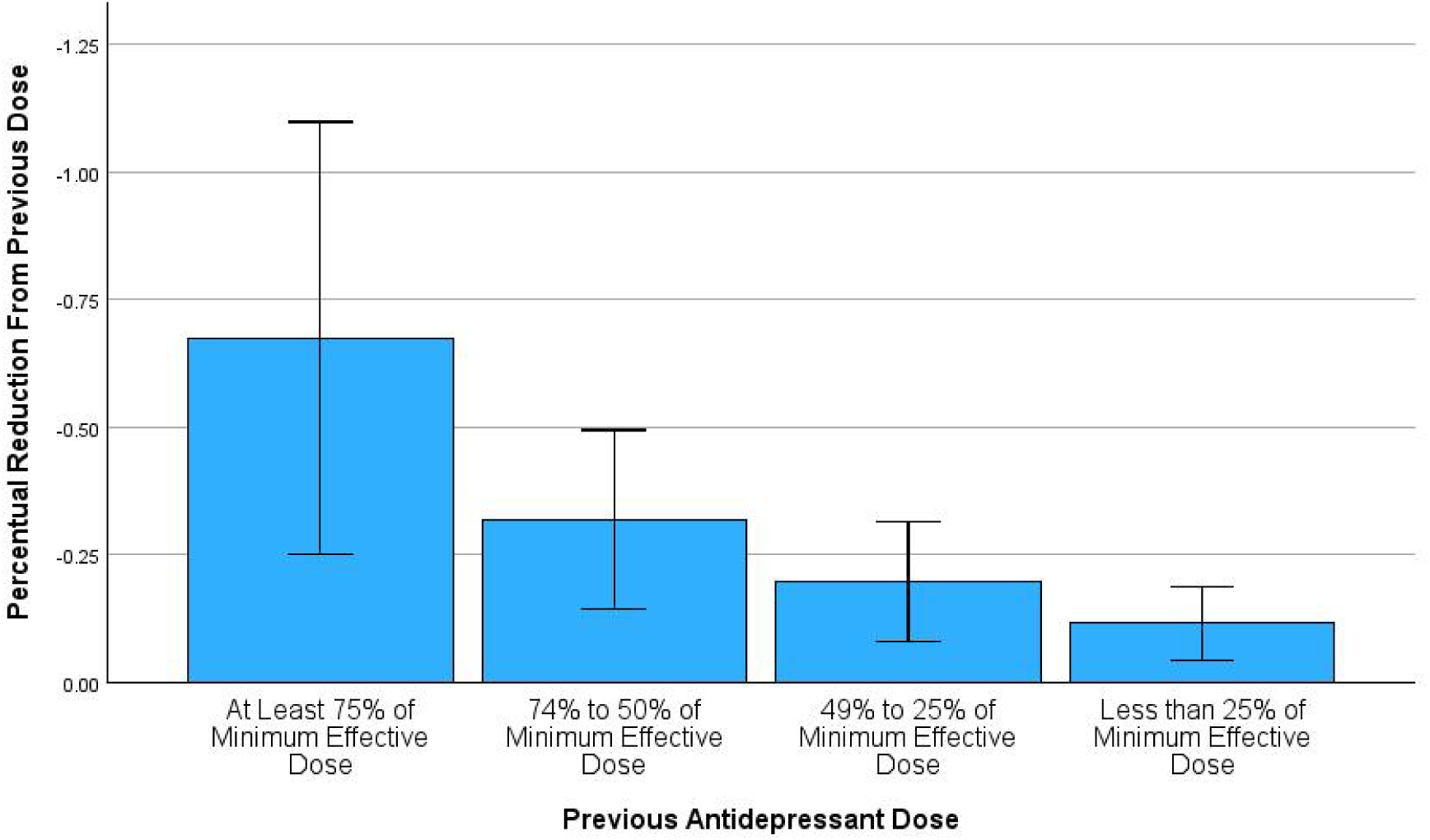
Average percentual reductions from the previous dose for different thresholds relative to the minimum effective dose across intervals. Error bars indicate ±1 standard deviation.

Figure 2 shows the DESS point and change scores over time. Across all three DESS measures, mean point scores were highest at baseline (week 0) and lowest at week 26. The largest intra-individual changes score over time, both negative and thus indicating mean decreases, occurred between baseline and week 2, followed by change from week 16 to 26. Mean intra-individual change during all other intervals were much smaller and fluctuated around zero, but with substantial interindividual variability, indicating that some participants experienced substantial symptom decreases, and others substantial symptom increases over time. The markedly increased mean DESS total score at baseline was largely due to three outliers with very high scores of 42, 47, and 58 points. The first was a middle-aged woman who indicated frequent and increased alcohol consumption, the second was an older woman with hypothyroidism treated with levothyroxine, and the third was a young woman with massive underweight (BMI of 16.1) requiring nutrition counselling. Correlations between the DESS scales at all time points and with the WHO-QoL-Bref at baseline are shown in the supplement. Noteworthy, at baseline the correlations between DESS measures and WHO-QoL-Bref were all statistically significant and moderate to large, ranging from r=-0.36 between DESS neurosensory and WHO-QoL-Bref mental wellbeing to r=-0.73 between DESS affective and WHO-QoL-Bref physical health, meaning that higher DESS baseline scores correlated with lower health-related quality of life.

**Figure 2:**
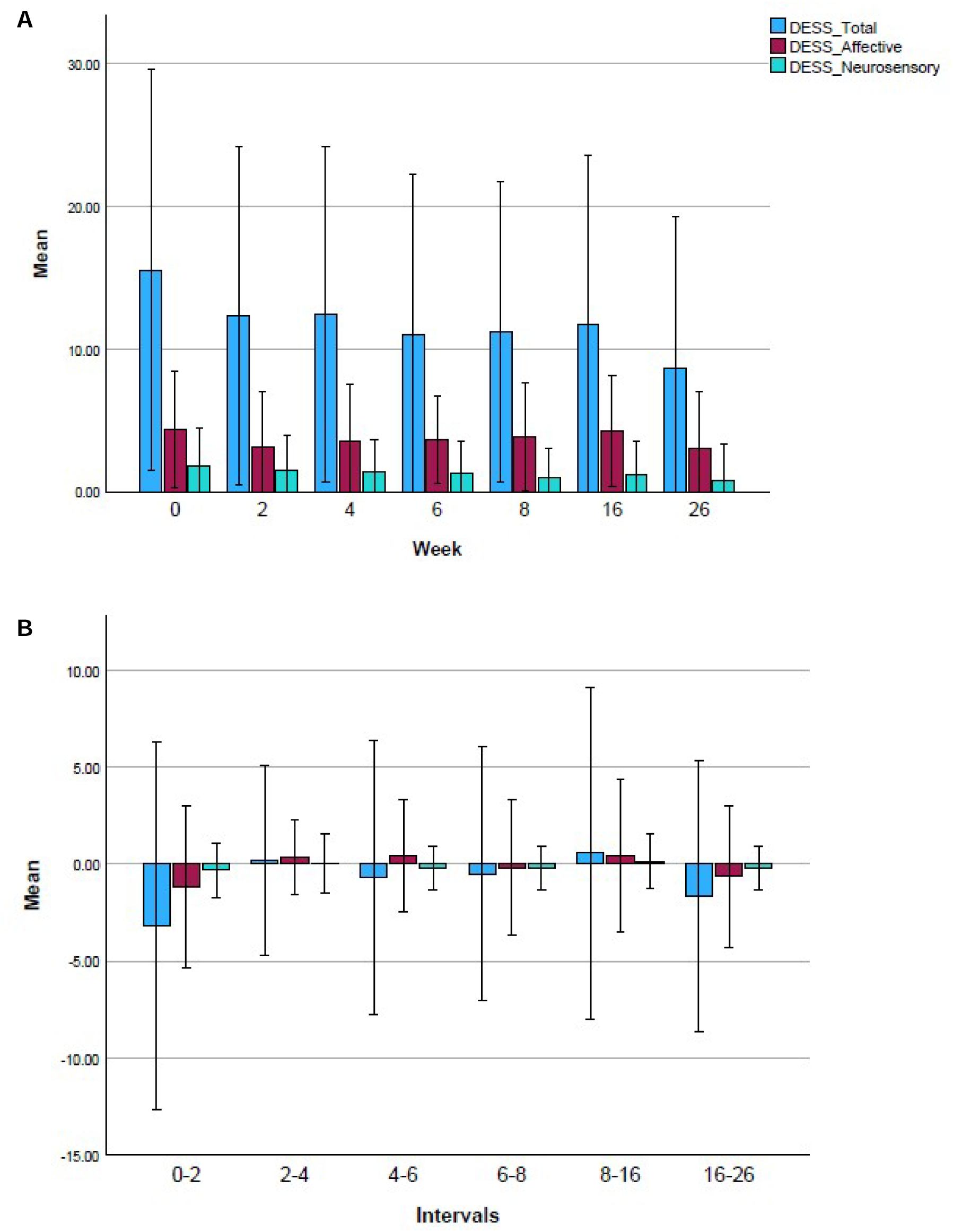
DESS point scores (graph A) and change scores (graph B) over time. Error bars indicate ±1 standard deviation.

We then examined whether this inter-individual variability in intra-individual symptom change was associated with concurrent changes in antidepressant dosage over time (see Table 2). The results show that intra-individual change in DESS scores was associated with distinct change in dosage. For DESS total, reductions below 75% of the minimum effective dose were associated with significantly higher change scores compared to intervals without dose reductions across all models, even when fully adjusted for various potential confounders (Model 3: beta=2.58, p<0.001). A similar pattern was observed for neurosensory symptoms (Model 3: beta=0.46, p<0.01) and affective symptoms (Model 3: beta=0.92, p<0.001). These differences correspond to standardized mean differences of 0.34 for DESS total, 0.35 for DESS neurosensory, and 0.27 for DESS affective. In contrast, intra-individual symptom change contingent on dose reductions above 75% of the minimum effective dose did not differ significantly from intervals when no dose reductions occurred and the differences were close to zero in the fully adjusted Models 3 (all beta<0.05). Overall, dose reductions below 75% of the minimum effective dose were associated with concurrent intra-individual symptom increase, whereas both reductions above 75% of the minimum effective dose and intervals with no dose reductions were associated with intra-individual symptom decrease.

**Table 2:** Repeated intra-individual change in DESS scores in association with concurrent change in dosage between adjacent time points measured at six intervals over 26 weeks.

| Outcome | Predictor | Model 1<br>Mean (95%-CI) | beta | Model 2<br>Mean (95%-CI) | beta | Model 3<br>Mean (95%-CI) | beta |
| --- | --- | --- | --- | --- | --- | --- | --- |
| DESS Total | Reduction below 75% of minimum effective dose | 0.72<br>(0.46 to 0.98) | 2.35*** | 0.89<br>(0.59 to 1.20) | 2.52*** | 0.84<br>(0.52 to 1.16) | 2.58*** |
|  | Reduction above 75% of minimum effective dose | -1.35<br>(-1.76 to -0.94) | 0.28 | -1.06<br>(-1.51 to -0.62) | 0.57* | -1.75<br>(-2.22 to -1.28) | -0.02 |
|  | No dose reduction | -1.63<br>(-1.82 to -1.44) | Ref. | -1.63<br>(-1.85 to -1.41) | Ref. | -1.74<br>(-1.99 to -1.48) | Ref. |
| DESS Neurosensory | Reduction below 75% of minimum effective dose | 0.16<br>(-0.10 to 0.42) | 0.42* | 0.19<br>(-0.11 to 0.49) | 0.44** | 0.18<br>(-0.14 to 0.50) | 0.46** |
|  | Reduction above 75% of minimum effective dose | -0.30<br>(-0.71 to 0.10) | -0.04 | -0.23<br>(-0.68 to 0.21) | 0.01 | -0.32<br>(-0.79 to 0.15) | -0.04 |
|  | No dose reduction | -0.26<br>(-0.45 to -0.07) | Ref. | -0.25<br>(-0.46 to -0.03) | Ref. | -0.28<br>(-0.53 to -0.03) | Ref. |
| DESS Affective | Reduction below 75% of minimum effective dose | 0.47<br>(0.21 to 0.73) | 0.91*** | 0.50<br>(0.20 to 0.81) | 0.98*** | 0.39<br>(0.07 to 0.71) | 0.92*** |
|  | Reduction above 75% of minimum effective dose | -0.17<br>(-0.58 to 0.23) | 0.27 | -0.32<br>(-0.77 to 0.12) | 0.15 | -0.55<br>(-1.02 to -0.08) | -0.02 |
|  | No dose reduction | -0.44<br>(-0.63 to -0.25) | Ref. | -0.47<br>(-0.69 to -0.25) | Ref. | -0.53<br>(-0.77 to -0.28) | Ref. |

Note:
\* p<0.05; \*\* p<0.01; \*\*\* p<0.001
Model 1: unadjusted
Model 2: controlling for drug class, standardized dosage at baseline, duration of antidepressant use, and DESS baseline score
Model 3: controlling for drug class, standardized dosage at baseline, duration of antidepressant use, DESS baseline score, age, gender, treatment indication, physical health at baseline, and mental wellbeing at baseline

To estimate the risk of pharmacological withdrawal reactions we used a categorical variable of intraindividual change in DESS total as outcome variable (clinically relevant increase of at least 4 points vs. less or no increase). Table 3 shows that clinically relevant increases in DESS total occurred significantly more often when the dose was reduced below 75% of the minimum effective dose compared to intervals without dose reduction (Model 3: OR=3.2; 1.4 to 7.4), whereas reductions above 75% of the minimum effective dose did not differ from no reductions (Model 3: OR=0.6; 0.2 to 2.2). In the fully adjusted analysis (Model 3), the estimated average risk of a clinically relevant intra-individual increase in DESS total was 33% during intervals with dose reductions below 75% of the minimum effective dose, as compared to 8% during intervals with dose reductions above 75% of the minimum effective dose and 13% during intervals with no dose reduction. Using the average rate during intervals without dose reductions as control, roughly 60% of the risk of clinically relevant increases in DESS total is likely attributable to pharmacological drug effects and conversely 40% of the risk is due to other factors (13/33=0.394).

**Table 3:** Estimated frequency of clinically relevant intra-individual increases in DESS total score in association with concurrent changes in dosage between adjacent time points measured at six intervals over 26 weeks.

| Predictor | Model 1 |  | Model 2 |  | Model 3 |  |
| --- | --- | --- | --- | --- | --- | --- |
|  | Rate | OR (95%-CI) | Rate | OR (95%-CI) | Rate | OR (95%-CI) |
| Reduction below 75% of minimum effective dose | 31% | 2.23 (1.05 to 4.72) | 35% | 2.51 (1.15 to 5.49) | 33% | 3.19 (1.37 to 7.40) |
| Reduction above 75% of minimum effective dose | 17% | 1.04 (0.32 to 3.43) | 14% | 0.74 (0.22 to 2.56) | 8% | 0.60 (0.17 to 2.18) |
| No dose reduction | 17% | Ref. | 17% | Ref. | 13% | Ref. |

*Note:*
Model 1: unadjusted
Model 2: controlling for drug class, standardized dosage at baseline, duration of antidepressant use, and DESS baseline score
Model 3: controlling for drug class, standardized dosage at baseline, duration of antidepressant use, DESS baseline score, age, gender, treatment indication, physical health at baseline, and mental wellbeing at baseline

By week 26, during at least one interval with a dose reduction below 75% of the minimum effective dose, n=7 participants (22%) had experienced a mild or subclinical increase in DESS total (1-3 points), n=11 (34%) had experienced a moderate increase (4-7 points), and n=6 (19%) had experienced a severe increase (8 or more points). Thus, 53% of all participants had experienced at least one clinically relevant increase in DESS total score contingent on a dose reduction below 75% of the minimum effective dose. If, as derived above, 60% of the risk for a clinically relevant increase in DESS total contingent on dose reduction below 75% of the minimum effective dose is due to pharmacological withdrawal effects, then the adjusted incidence rate of a clinically relevant withdrawal reaction is 32% and the adjusted incidence rate of a severe withdrawal reaction is 11%.

## Discussion

### Summary of findings

In this prospective longitudinal cohort study of primary care patients followed over 26 weeks we found that differences in intra-individual change in withdrawal symptoms assessed with the DESS were significantly related to concurrent changes in dosage. Longitudinal repeated-measure analyses showed that during intervals when the dose was reduced below 75% of the minimum effective dose, DESS change scores increased on average, whereas they declined when the dose was reduced above 75% of the minimum effective dose or when no dose reduction occurred. Moreover, we found that during intervals when the dose was reduced below 75% of the minimum effective dose, clinically relevant intra-individual increases in DESS scores occurred 33% of the time, while the rate was 13% when no dose reduction occurred. Based on this we calculated that about 60% of the risk of clinically relevant intra-individual increases in DESS scores contingent on dose reductions below 75% of the minimum effective dose is due to pharmacological withdrawal effects. By the end of the 26-week observation period, 53% of the sample had experienced at least once a clinically relevant increase in DESS contingent on a dose reduction below 75% of the minimum effective dose. Assuming that 60% of the risk is attributable to pharmacological withdrawal effects, the adjusted incidence rate of clinically relevant withdrawal reactions is 32% and the adjusted incidence rate of severe withdrawal reactions is 11%.

### Interpretation and contextualization

The present study helps to advance the ongoing scientific controversy over the incidence and clinical relevance of pharmacological withdrawal reactions [16,17,28]. We found that slightly over half of patients (53%) experience a clinically relevant symptom increase when coming off antidepressants, in agreement the available evidence from clinical and observational studies with a systematic assessment of antidepressant withdrawal [17,33]. However, symptom increases are not exclusively due to pharmacological withdrawal effects. Meta-analyses of randomized controlled trials reported an incidence rate of roughly 17% for “withdrawal syndromes” occurring in antidepressant continuation groups [34]. In correspondence, our analysis showed that about 60% of the risk of clinically relevant increases in DESS scores contingent on dose reductions below 75% of the minimum effective dose is likely due to pharmacological withdrawal effects and conversely 40% of the risk may be attributable to non-pharmacological effects such as external stressors or random symptom fluctuations due to concomitant health problems. However, this calculation does not take into account the possibility of delayed onset withdrawal which may have contaminated the symptom change during 2-week intervals without dose reduction. This possibility means that pharmacological effects may have explained more than 60% of the increase in DESS score. Adjusting the cumulative risk of clinically relevant increases DESS scores contingent on dose reduction below 75% of the minimum effective dose produces an adjusted incidence rate of 32% for pharmacological withdrawal reactions. Noteworthy, according to the meta-analysis of clinical trials by Zhang et al. [34], the pooled rate of antidepressant withdrawal syndromes is 44.4% in antidepressant discontinuation groups and 16.6% in antidepressant continuation groups, thus producing an adjusted incidence rate of roughly 28% (44.4% - 16.6%). This estimate is comparable to our adjusted incidence rate of 32% for moderate to severe pharmacological withdrawal reactions. Our 60% estimate of the pharmacological effect may thus be used as a conservative means to adjust incidence rates of withdrawal syndromes based on changes in DESS total scores derived from clinical trials, observational studies and patient surveys lacking adequate control groups.

From a clinical perspective it is important to stress that the mean duration of antidepressant use in our study was 42 months (3.5 years), with a standard deviation of 47 months. This finding is consistent with representative national Swiss data from statutory health insurances, showing that over half of current antidepressant users are on their drug for over two years and that the rate of long-term users is steadily increasing [3]. Most participants in our study had not attempted to discontinue their medication before, raising questions about the appropriateness of ongoing treatment monitoring and drug reviews in primary care [35,36]. We also note that two-thirds of participants in our study had not received a tapering plan from their treating physician, which further indicates that adherence to clinical guidelines may be inadequate, especially when it comes to treatment discontinuation and deprescribing. Finally, our results show that dose reductions in the therapeutic range are mostly unproblematic, but when the dose is reduced below the minimum effective dose, the risk of pharmacological withdrawal reactions is significantly increased even though average dose reductions became increasingly smaller. An individualised long-term tapering plan below 75% of the minimum effective dose is therefore needed when clinically relevant withdrawal symptoms emerge. Physicians should be advised that very small reductions are required in the lowest dose-range for some patients and that in some cases, tapering may take several months and should be monitored regularly [20,37].

### Strength and limitations

Our study has several strengths, including a prospective cohort study design, longitudinal repeated measures and systematic assessment of antidepressant dosage and withdrawal symptoms, and long observation period of 26 weeks. However, we also acknowledge the following major limitations. We achieved to recruit only 38 participants, of which 32 completed the study, instead of the 400 participants originally planned in the study protocol. This is because we did not anticipate that the participating GPs would recruit so few participants. Unfortunately, this is a well-known problem in research involving GPs, and poor recruitment is frequently reported in both clinical and observational studies in primary care, which often leads to study discontinuation [38,39]. Various factors affect poor recruitment, among which GPs high workload and lack of resources arguably are the most influential [39,40]. Various analyses originally planned, such as detection of predictor and moderator variables of withdrawal reactions, were not possible. Nevertheless, due to the repeated measurements, statistical power was adequate to assess withdrawal reactions of at least moderate severity (see the supplement for more information). Moreover, although the sample was small, it was broadly representative of the Swiss adult population of antidepressant users in terms of gender, age, treatment duration, and drug class [3,41].

## Conclusions

Primary care patients are often on antidepressants for many years. GPs should conduct regular drug reviews and evaluate, whether such long-term antidepressant therapy is indicated and in agreement with clinical practice guidelines to avoid unnecessary and potentially harmful treatment. Clinically relevant symptom increases during antidepressant tapering most likely emerge following dose reduction below 75% of the minimum effective dose. About 60% of the risk of clinically relevant intra-individual symptom increases is attributable to pharmacological withdrawal effects, thus the estimated incidence rate for a clinically relevant withdrawal reaction is 32%, and for a severe withdrawal reaction it is 11%. Practitioners should be aware that even small reductions in the dose range well below the minimum effective dose can trigger withdrawal reactions, and in such cases, personalized tapering plans and close monitoring of withdrawal symptoms are recommended.

## Ethics approval and consent to participate

The study was approved by the Cantonal Ethic Committee of Zurich on September 27, 2022, reference number 2022-01498. All study participants gave written informed consent.

## Consent for publication

All authors approved the final version of the manuscript.

## Data Availability

Data and statistical code are freely available on the Open Science Framework, https://osf.io/aqmyt/overview

## Acknowledgements

We want to thank all contributing physicians who helped to recruit study participants.

