## Supplement for "Incidence of antidepressant withdrawal reactions: A prospective longitudinal cohort study in primary care patients"

**Supplementary materials**

1. **Additional recruitment strategies**

Since the recruitment strategies prespecified in the study protocol turned out to be insufficient, additional recruitment strategies were introduced. After randomly selecting 400 GPs from the database of the Swiss Medical Association, an invitation by email was also sent to all 727 members of the Swiss GP research database FIRE, followed by an invitation to the 198 members of the Zurich GP network mediX and 115 physicians contacted personally during quality-circle meetings organized by the PizolCare network in the Sarganserland region. In addition, 750 GPs identified through online directories were contacted by email. Due to the persistently low number of referrals from GPs, recruitment was expanded in 2023 to include outpatient psychiatrists working in primary care practices. The study was presented in five cantonal psychiatric outpatient clinics in the canton of St. Gallen, reaching 66 psychiatrists. Finally, 80 outpatient psychiatrists working in private practice identified through online directories were contacted by telephone and email. In total, over 2000 physicians in the German-speaking part of Switzerland were contacted and invited to recruit participants for study inclusion.

2. **Sample size calculation and post-hoc power analysis**

A priori power analysis based on a repeated-measures MANCOVA indicated that a total sample of 306 participants would be required to detect a small within-person effect size (f = 0.1) over 4 measurements across 3 groups (abrupt discontinuation, fast taper, slow taper) with a power of 0.95 at an alpha level of 0.05. For a medium effect size (f=0.25) a total of 54 participants would have been sufficient. Those calculations were based on our initial plan to analyse the acute withdrawal phase separately, assuming that most participants would discontinue their antidepressant within 8 weeks. Unfortunately, it was impossible to recruit that many participants because it was very difficult to find GPs willing to participate in the study and because the participating GPs repeatedly asserted that they had not seen eligible patients. Moreover, it turned out that many participants used slow tapers, thus not reducing their dose below 75% of the minimal therapeutic dose within the first eight weeks. Post-hoc power analysis focusing on repeated intraindividual changes up to week 26 (6 measurements) showed that with 32 participants we achieved a power of 0.83 to find a medium effect size of f=0.25.

3. **Descriptive statistics**

Mean DESS scores at different time points and intra-individual change during intervals

| **Time Points and Intervals** | **DESS Total** |  | **DESS Neurosensory** |  | **DESS Affective** |  |
| --- | --- | --- | --- | --- | --- | --- |
|  | Mean (SD) | Range | Mean (SD) | Range | Mean (SD) | Range |
| Week 0 | 15.48 (14.03) | 1 to 58 | 1.81 (2.69) | 0 to 11 | 4.32 (4.09) | 0 to 17 |
| Change to week 2 | -3.19 (9.44) | -19 to 22 | -0.32 (1.42) | -3 to 2 | -1.19 (4.16) | -9 to 14 |
| Week 2 | 12.29 (11.82) | 0 to 44 | 1.48 (2.42) | 0 to 10 | 3.13 (3.84) | 0 to 17 |
| Change to week 4 | 0.19 (4.88) | -14 to 10 | 0.0 (1.53) | -3 to 5 | 0.32 (1.94) | -5 to 6 |
| Week 4 | 12.41 (11.72) | 0 to 47 | 1.44 (2.17) | 0 to 9 | 3.56 (3.93) | 0 to 16 |
| Change to week 6 | -0.71 (7.09) | -19 to 15 | -0.19 (1.14) | -4 to 2 | 0.45 (2.90) | -7 to 7 |
| Week 6 | 11.00 (11.17) | 0 to 48 | 1.29 (2.19) | 0 to 10 | 3.61 (3.07) | 0 to 9 |
| Change to week 8 | -0.53 (6.55) | -19 to 16 | -0.22 (1.13) | -2 to 4 | -0.19 (3.48) | -7 to 10 |
| Week 8 | 11.19 (10.56) | 0 to 41 | 1.03 (1.94) | 0 to 9 | 3.81 (3.76) | 0 to 14 |
| Change to week 16 | 0.56 (8.57) | -20 to 21 | 0.13 (1.41) | -3 to 5 | 0.44 (3.93) | -10 to 11 |
| Week 16 | 11.75 (11.74) | 0 to 50 | 1.16 (2.36) | 0 to 11 | 4.25 (3.84) | 0 to 15 |
| Change to week 26 | -1.67 (6.98) | -16 to 18 | -0.23 (1.14) | -3 to 3 | -0.63 (3.65) | -10 to 10 |
| Week 26 | 8.60 (10.69) | 0 to 51 | 0.77 (2.57) | 0 to 14 | 3.03 (4.00) | 0 to 18 |

4. **Preliminary analysis**

GEE Model: Repeated intraindividual change in DESS total over time (outcome variable) in association with concurrent dosage relative to 75% of minimal therapeutic dose (0 “below” vs. 1 “above”) and dose change (0 “no reduction” vs. 1 “reduction”); main effects only

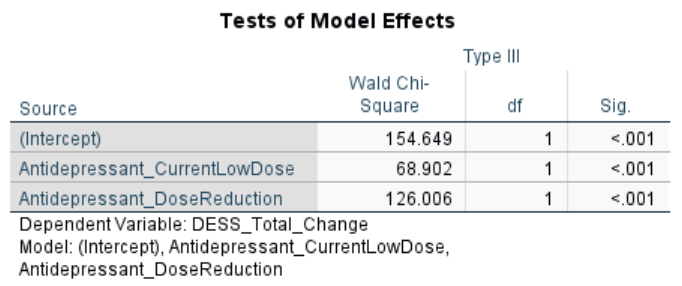

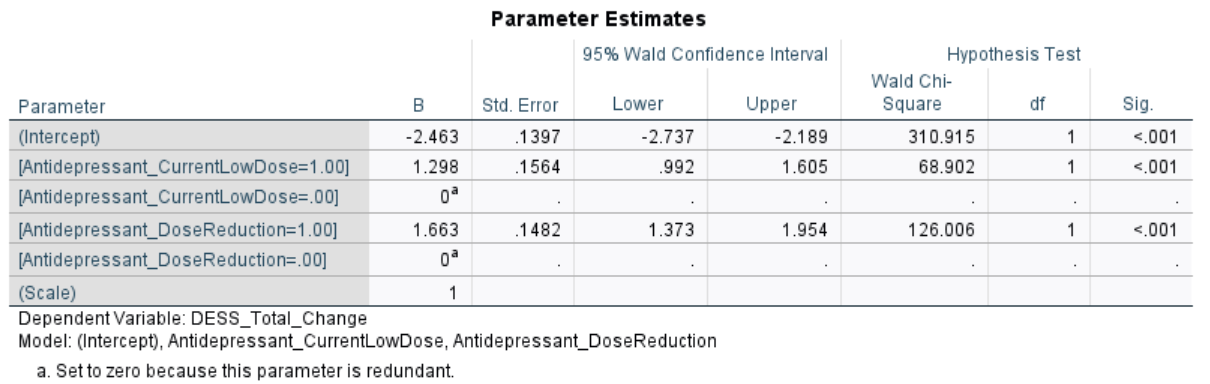

GEE Model: Repeated intraindividual change in DESS total over time (outcome variable) in association with concurrent dosage relative to 75% of minimal therapeutic dose (0 “below” vs. 1 “above”) and dose change (0 “no reduction” vs. 1 “reduction”); main effects and interaction effect

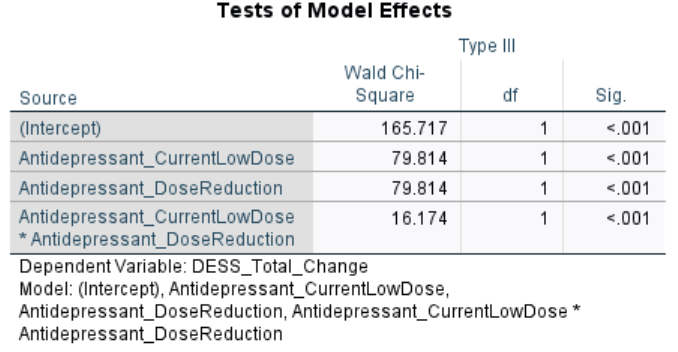

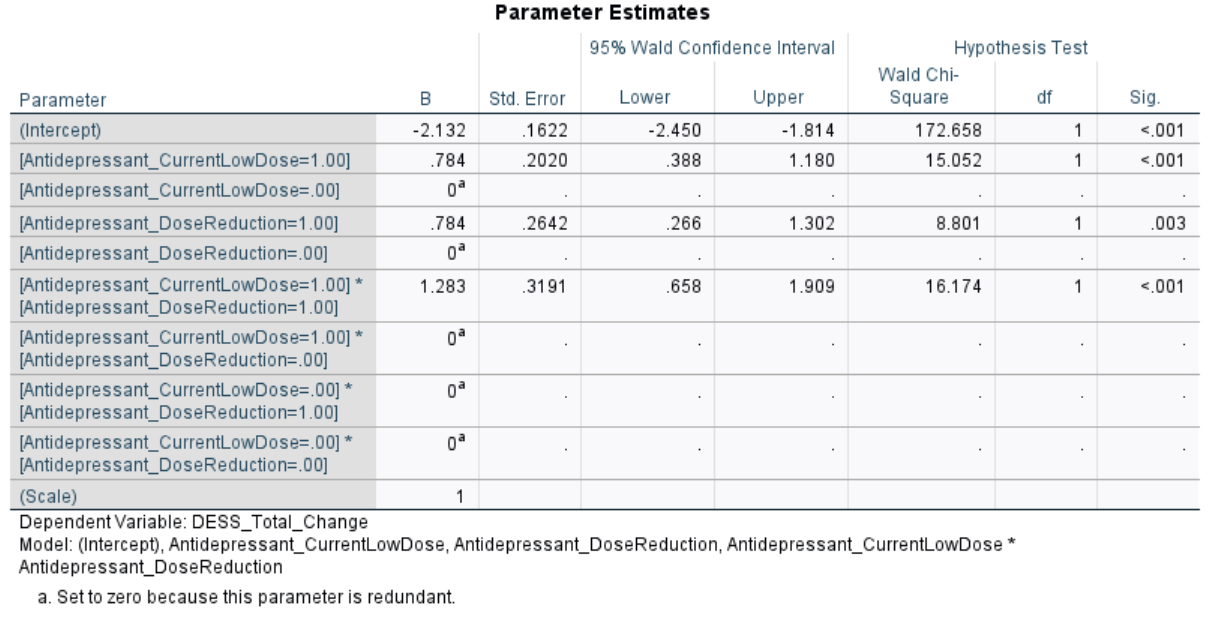

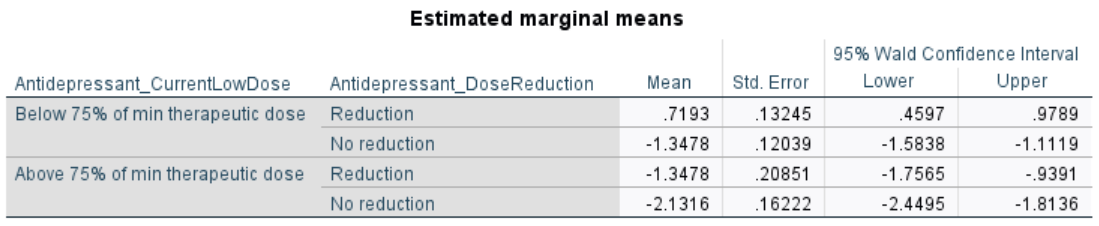

GEE Model: Repeated intraindividual change in DESS total over time (outcome variable) in association with dose reductions relative to minimal therapeutic dose; comparison of different thresholds (the smaller the values, the better the model fit)

Dosage changes above vs. below 75% of minimal therapeutic dose

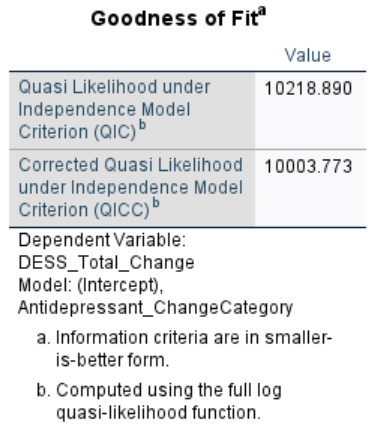

Dosage changes above vs. below 100% of minimal therapeutic dose

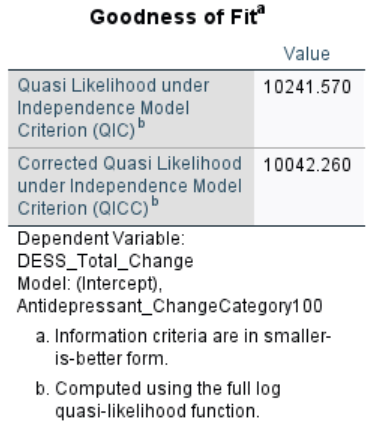

Dosage changes above vs. below 125% of minimal therapeutic dose

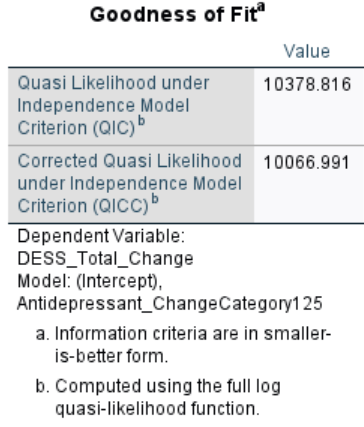

5. **Correlations**

Pearson correlation between DESS and WHO-QoL-Bref measures at week 0 (baseline)

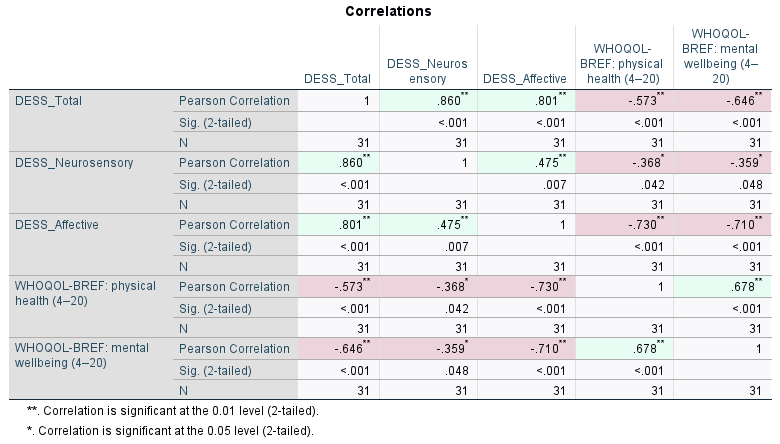

Pearson correlation between DESS measures at week 2

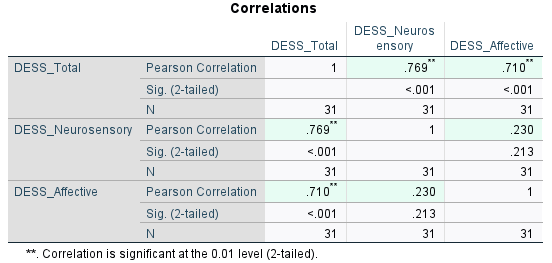

Pearson correlation between DESS measures at week 4

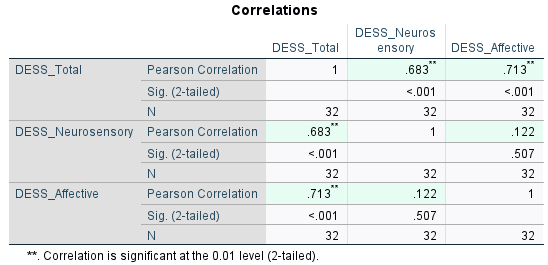

Pearson correlation between DESS measures at week 6

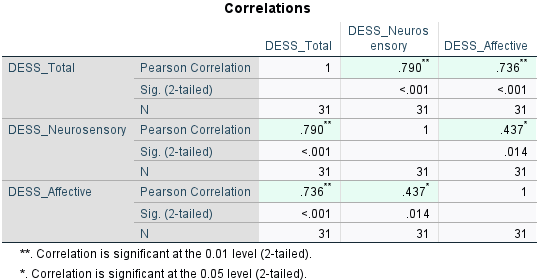

Pearson correlation between DESS measures at week 8

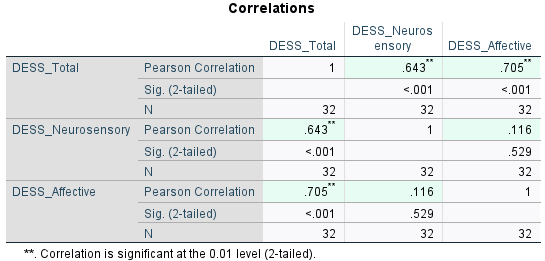

Pearson correlation between DESS measures at week 16

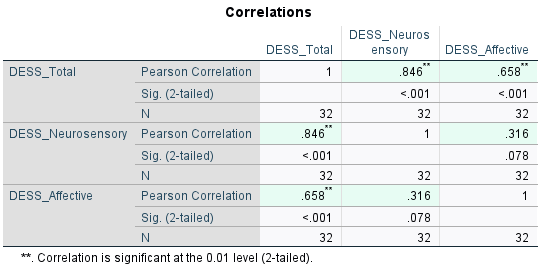

Pearson correlation between DESS measures at week 26

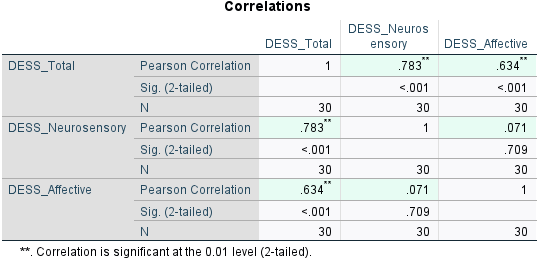
